# Data sharing in the age of predictive psychiatry: An adolescent perspective

**DOI:** 10.1101/2021.08.25.21262234

**Authors:** Gabriela Pavarini, Aleksandra Yosifova, Keying Wang, Benjamin Wilcox, Nastja Tomat, Jessica Lorimer, Lasara Kariyawasam, Leya George, Sonia Alí, Ilina Singh

## Abstract

**Background:** Advances in genetics and digital phenotyping in psychiatry have given rise to testing services targeting young people, which claim to predict psychiatric outcomes before difficulties emerge. These services raise several ethical challenges surrounding data sharing and information privacy.

**Objectives:** This study aimed to investigate young people’s interest in predictive testing for mental health challenges, and their attitudes towards sharing biological, psychosocial and digital data for such purpose.

**Methods:** Eighty UK adolescents aged 16-18 participated in the study. Participants took part in a digital role-play where they played the role of clients of a fictional predictive psychiatry company and chose what sources of personal data they wished to provide for a risk assessment. After the role-play, participants reflected upon their choices during a peer-led interview.

**Findings:** Participants saw multiple benefits in predictive testing services, but were highly selective with regards to type the data they were willing to share. Largely due to privacy concerns, digital data sources such as social media or Google search history were less likely to be shared than psychosocial and biological data, including school grades and one’s DNA. Participants were particularly reluctant to share digital data with schools (but less so with health systems).

**Conclusions:** Emerging predictive psychiatric services are valued by young people; however, these services must consider privacy vs. utility trade-offs from the perspective of different stakeholders, including adolescents.

**Clinical implications:** Respecting adolescents’ need for transparency, privacy and choice in the age of digital phenotyping is critical to the responsible implementation of predictive psychiatric services.

## BACKGROUND

Psychiatry has traditionally relied on behavioural observations, genomics, and neuroscience to draw conclusions about the aetiology of mental health challenges and build predictive risk models [1]. However, over the past two decades, as digital data have become increasingly ubiquitous, other data sources have emerged as relevant for early detection of psychiatric problems. These include activity data, sleep patterns, speech features, social media interaction and typing speed, all of which can be collected via smartphones and wearable sensors [2,3]. Coupled with appropriate analytical methods, these multiple sources of data might be used to build models capable of predicting psychiatric outcomes for individual patients, moving beyond the investigation of group-level statistical associations [4,5]

Such advances impact young people, particularly minors, because young people are heavy users of digital technologies [6]. They are also likely to be the main targets for early screening and early intervention programmes in mental health based on big data technologies [5,7]. Indeed, even though research is still in its infancy, a number of private companies have already started offering predictive psychiatric testing services. Social Sentinel (https://www.socialsentinel.com/) scans digital signals to detect safety and security threats in school, including self-harm; Steer (www.steer.global) and Impero (https://www.imperosoftware.com/) offer affective and emotional tracking for early identification of mental health risk; 23andme (www.23andme.com), which largely targets young people, offers predictive genetic testing for Alzheimer’s disease. There is also enthusiasm for the possibility of linking/sharing data between schools and health systems to support research and intervention in adolescent mental health [8] and for health systems and schools to use social media data to monitor mental health risk [9].

Scientific advances in predictive mental health, alongside the increasing availability of commercial testing services, have sparked significant controversy and debate over potential ethical and social implications [10–12]. For instance, researchers have questioned the current utility of predictive tests in psychiatry, given their limited predictive power [13,14]. Others have pointed to the unique sensitivity of mental health data, which may not only be used for accessing health and social services, but also influence criminal justice proceedings (e.g., sentence mitigation), and might attract stigma, discrimination and forced treatment [15]. Associated issues surrounding who owns and who should be granted access to mental health data have also been raised [12,16]. In the context of minors, researchers have debated who has the right to manage a child’s risk, and raised privacy concerns regarding data sharing [11,17,18].

Before predictive mental health services are widely implemented, it is essential to understand the extent to which young people value these services, and their attitudes and preferences in relation to sharing different sources of personal data for such purpose. Empirical ethics research so far has largely focused on predictive testing based on genetic information, but not a combination of data sources, especially digital. Small-scale studies suggest that young people’s interest in learning about their genetic susceptibility for psychosis is low among nonclinical populations [19] and high among clinical high-risk participants [20]. Yet, high-risk participants expressed worries surrounding stigma, data privacy and potential psychological harm of genetic risk information for psychosis [21]. Similar concerns were expressed by grandchildren of people with late-onset Alzheimer’s disease with regards to testing for a susceptibility gene [22].

It is unknown, however, whether existing evidence related to psychosis and Alzheimer’s holds for attitudes to risk prediction for other psychiatric or neurological difficulties. It is also unclear whether risk predictions based on genomic data will map onto prediction based on diverse data sources, including psychosocial and particularly digital data. There is research indicating that adolescents value learning about themselves and their health [23]. However, recent findings suggest that adolescents view their Internet use as a personal activity and are concerned about the privacy of their digital information [24].

## OBJECTIVE

Our study aimed to investigate adolescents’ interest in services that predict risk of mental health challenges, and their preferences and attitudes towards sharing personal biological, psychosocial, and behavioural data for this purpose. We investigated the following questions:

a. Would young people want to take a predictive test for mental health challenges, and if so, which conditions and what motivates them to do so?
b. What types of data (biological, psychosocial, digital) are young people willing to share for a predictive test, and how do they make this choice?
c. What are young people’s attitudes towards sharing data with schools and health systems for early identification of mental health challenges?

## METHODS

The study was pre-registered at the Open Science Framework and all materials are available at https://osf.io/cwjx4/. Ethics approval was granted by the University of Oxford Medical Sciences Interdivisional Research Ethics Committee (Ref: R38020/RE001). We recruited young people aged 16-18 years from schools in South East England. Parents were notified about the study, and participants provided informed consent before participating in the session. A sample size of 80 participants was pre-determined based on a previous report of young people’s attitudes towards testing for psychiatric conditions [19].

### Procedure

To engage young people, we used digital role-play, combined with peer-led interviews. Our digital role-play, titled *What Lies Ahead?*, simulated the experience of being offered a predictive service for risk of specific mental health conditions. In a validation study, the scenario was shown to successfully immerse young people and elicit both authentic responses and reflective thinking [25]. Peer-led interviews were chosen because they have been found to be more comfortable and engaging for young people than adult-led interviews, as well as to improve the quality of collected data from young people [26].

Sessions took place at the participants’ schools, and in each session two randomly paired students were guided by a researcher (GP, JL, KW, LK or LG). Interview sessions took approximately one hour, and started with a brief icebreaker for the two participants to meet each other. *What Lies Ahead?* then collected data on participants’ choices around predictive mental health testing and provided an entry point for the peer-led interview, which acted as a platform for adolescents to articulate their attitudes and preferences.

### Youth Involvement

*What Lies Ahead?* was co-produced with the NeurOX Young People’s Advisory Group, a diverse group of 15-18-year-olds who share an interest in ethics and mental health (https://begoodeie.com/ypag/). This was done through a series of interactive group sessions, where young people co-produced the role-play concept, and gave input into the visuals and script. NeurOX YPAG members also co-led a pre-test of the role-play concept at a youth conference, and gave input into the interview guide. Part of the digital role-play was independently designed and produced by a group of five Work Experience students aged 16-18 years.

### Role-playing scenario: Signing up to a predictive test

Each participant independently completed the *What Lies Ahead?* digital role-play, on a computer or laptop. The role-play was presented from a first-person perspective (no avatar on the screen) and consisted of participants interacting with a staff member from “Future Forecast”, a predictive mental health company. The staff member presents the service and offers them the opportunity to sign up to learn their changes of facing mental health challenges in the future. Players are then asked to fill in forms on-screen to indicate whether they would like to take a test, and if so, for what challenges (e.g., depressed mood, anxiety) and what data they would be happy to provide for the assessment, including digital (e.g., social media, GPS), psychosocial (e.g., lifestyle information) and biological (e.g., hormone levels, DNA) (see **Table 1** for full list of items). Items were elaborated to reflect a range of mental health challenges and individual-level data sources used in psychiatric assessments and recent digital phenotyping research.

**Table 1.**
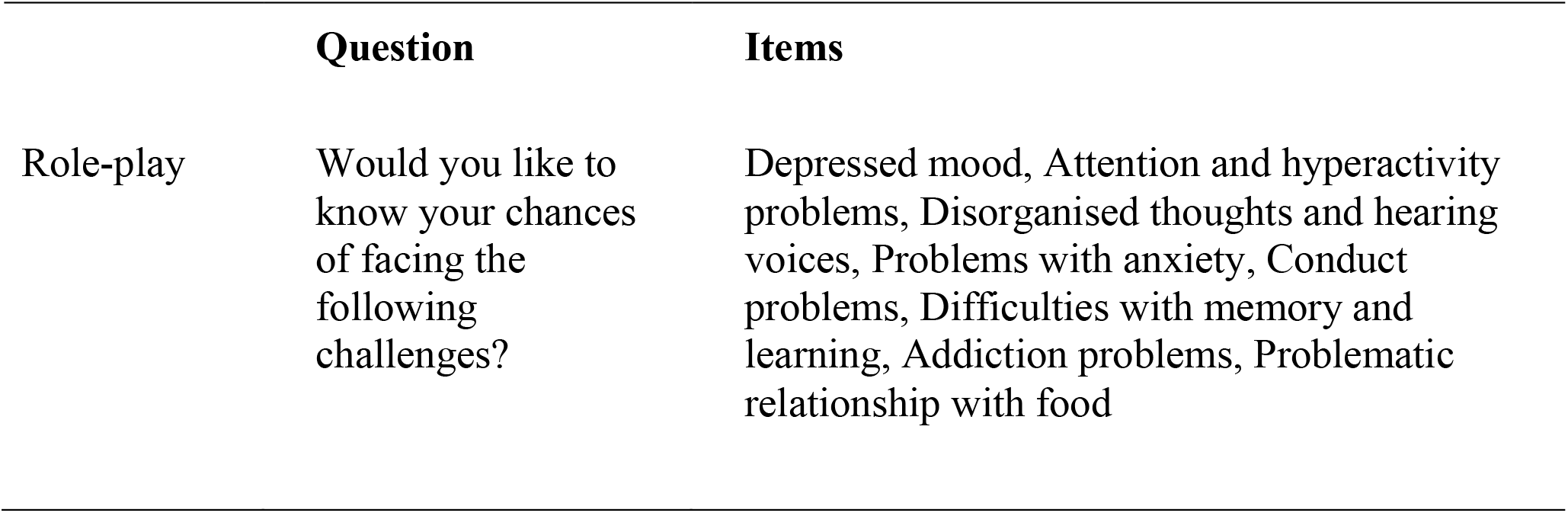
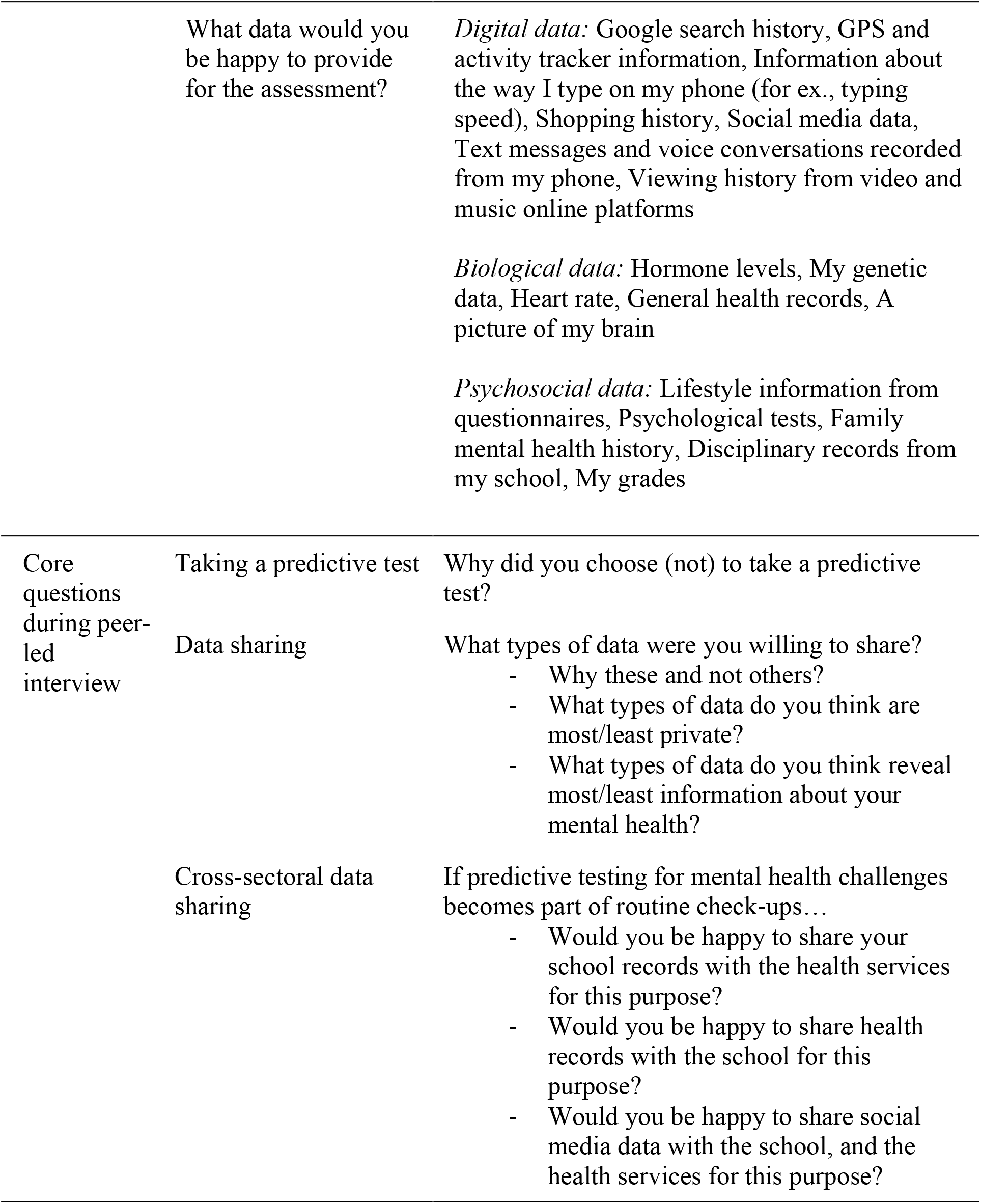
Role-play and interview questions used to measure interest in mental health predictive test, as well as preferences and attitudes towards data sharing

|  | Question | Items |
| --- | --- | --- |
| Role-play | Would you like to know your chances of facing the following challenges? | Depressed mood, Attention and hyperactivity problems, Disorganised thoughts and hearing voices, Problems with anxiety, Conduct problems, Difficulties with memory and learning, Addiction problems, Problematic relationship with food |
|  | What data would you be happy to provide for the assessment? | *Digital data:* Google search history, GPS and activity tracker information, Information about the way I type on my phone (for ex., typing speed), Shopping history, Social media data, Text messages and voice conversations recorded from my phone, Viewing history from video and music online platforms<br>*Biological data:* Hormone levels, My genetic data, Heart rate, General health records, A picture of my brain<br>*Psychosocial data:* Lifestyle information from questionnaires, Psychological tests, Family mental health history, Disciplinary records from my school, My grades |
| Core questions during peer-led interview | Taking a predictive test<br>Data sharing<br>Cross-sectoral data sharing | Why did you choose (not) to take a predictive test?<br>What types of data were you willing to share?<br>- Why these and not others?<br>- What types of data do you think are most/least private?<br>- What types of data do you think reveal most/least information about your mental health?<br>If predictive testing for mental health challenges becomes part of routine check-ups...<br>- Would you be happy to share your school records with the health services for this purpose?<br>- Would you be happy to share health records with the school for this purpose?<br>- Would you be happy to share social media data with the school, and the health services for this purpose? |

### Peer-led interview

After the role-play, participants took part in a qualitative peer-led interview, in which they took turns to ask and answer predefined questions to each other (drawing from a pile of flashcards). The questions prompted participants to justify and reflect on their choices during role-play. The questions covered their reasons to get a predictive test and for sharing different data sources. The interview guide also included questions about their preferences and attitudes with regards to sharing data across systems (which was not covered in the role-play). Namely, questions covered attitudes towards schools and public health services accessing health and school records, respectively, as well as social media data, to assess risk of mental health challenges. While participants led the interview, the researcher mediated the interaction, offered clarifications, and asked follow-up questions. Main questions are presented in **Table 1**. Interviews were audio-recorded and transcribed in full.

### Demographics and debrief

At the end of the session, participants filled in a brief demographic questionnaire. A short debriefing session followed, with the researcher emphasising that “Future Forecast” is a hypothetical company, and that predictive testing services as presented in the role-play are not currently available. Researchers also clarified any potential questions or concerns^1^.

### Data analysis

We used descriptive statistics to characterise participants’ interest in predictive testing for different conditions and their willingness to share different types of data (biological, psychosocial, and digital). Anonymised interview transcripts were analysed using the principles of thematic analysis, as guided by Braun and Clarke (2006). We developed separate thematic frameworks for the sections of the interview that referred to *interest in predictive testing, sharing personal data*, and *cross-sectoral data sharing*. The frameworks were based on the independent analysis of 25% of the transcripts by three different members of the research team; different interpretations of the data were discussed, and consensus was achieved on the main categories to be coded. We then followed an iterative process of coding the rest of the transcripts and refining themes to best reflect the core ideas expressed by participants. Participants were assigned pseudonyms; all names attached to quotes in the Findings section are pseudonyms.

## FINDINGS

There were 80 participants in the study, aged from 16 to 18 years (*M*age = 16.9; *SD* = 0.45), across six state schools. Participants were mainly females (59 females, 21 males) and were of different ethnicities (37% Asian or Asian British; 34% White; 19% Black, African or Black British; 10% mixed or other).

### Interest in predictive testing

Results from the digital role-play indicated that the majority of participants chose to take a predictive test for mental health challenges. Most participants chose to learn their risk for anxiety, learning difficulties and depression; the least chosen conditions were conduct problems and eating disorders (**Fig1**, top). Participants’ main expressed motivations to take the test are summarised in **Fig1**, alongside illustrative quotes. Two thirds of participants referred to the **relevance of particular mental health challenges to their past and present lives**. Relevant experiences included: awareness of being exposed to risk factors such as exam stress or family history of the condition; experience of similar challenges in the past; current experience of early signs of the condition; or more generally, a feeling that certain conditions are relevant to their daily lives. An equally common theme was **curiosity to learn about oneself and one’s future**. This included finding it interesting to learn about one’s mental health and particular conditions, and more generally expressing that the more information they could obtain, the better. A third theme, mentioned by about a fifth of the interviewees, was the notion that being aware of one’s risk of developing a certain mental health problem later in life could **support action readiness**. This included the ability to take action to prevent challenges and (less frequently) the chance to “prepare for it” from a psychological perspective. Only five participants expressed **no interest** in taking a test, either because they were “scared to find out” or generally uninterested.

**Fig1.**
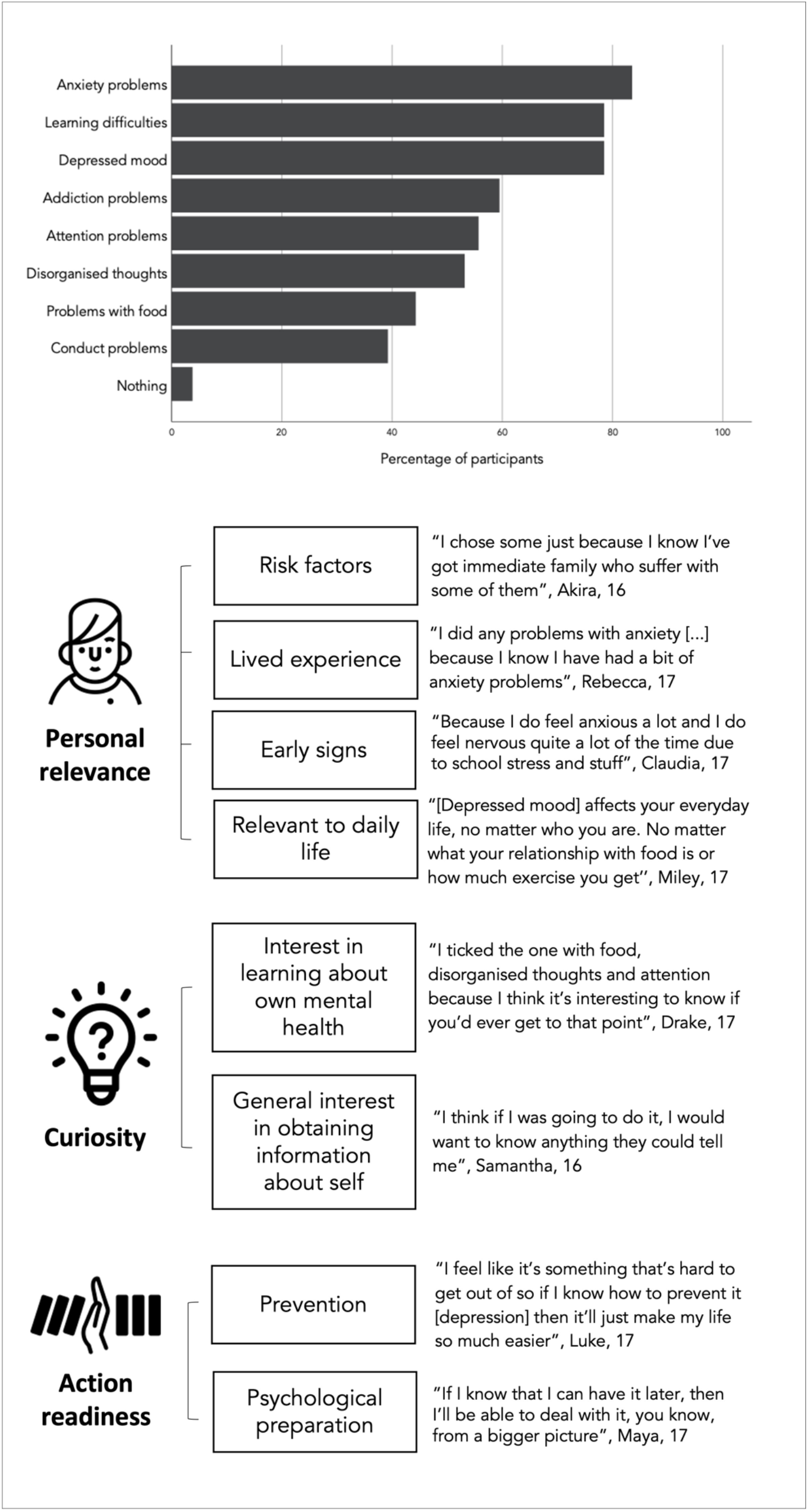
Percentage of participants interested in taking a predictive test for different mental health challenges (from digital role-play) and main reasons for taking a test (from interviews)

### Preferences and attitudes towards data sharing

Most participants were willing to share personal data for a predictive mental health test. However, participants’ willingness to share personal information depended on data type (**Fig2a**). While most participants were willing to share biological (66.5% on average) and psychosocial data (69.5% on average), only 30.7% of participants chose to share digital data sources. This average was even lower (26%) when excluding “typing patterns”. Two core themes were identified from participants’ justifications of data sharing choices: considerations around the **relevance of the data source** for predicting mental health issues and **privacy concerns** regarding disclosure. Data sources perceived as more useful for the test were more likely to be shared than data considered less relevant to the prediction. On the other hand, data sources considered more private were less likely to be shared than data considered less private (see **Fig2b** for quotes). Participants often made trade-off decisions, placing data sources across these two dimensions of relevance and privacy.

**Fig2.**
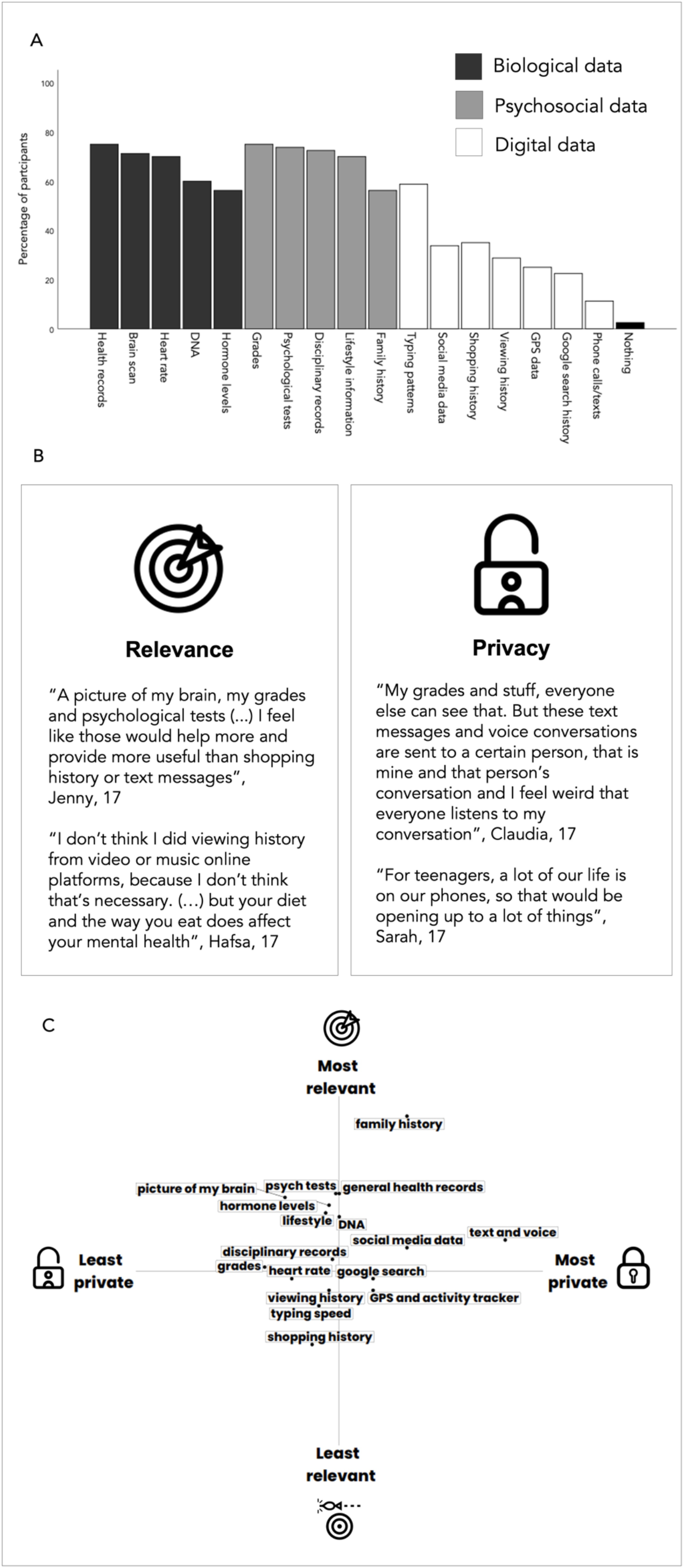
**A**. Percentage of participants who shared different sources of personal data for mental health predictive test (from role-play). **B**. Main reasons for sharing data (from interview). **C**. Indicative two-dimensional configuration of data sources according to perceived privacy and relevance. Axes indicate the frequency with which the data source was referred to by participants when asked to comment on data sources most/least private and most/least revealing of their mental health (from interview)

In **Fig2c** we present an indicative, two-dimensional configuration of types of data participants referred to as most and least private, and most and least relevant to a risk assessment. As illustrated, digital data sources such as Google search and GPS tended to be perceived as private; specific digital data sources perceived as not very private such as shopping history and typing speed tended to also be perceived as not as relevant. Psychosocial and biological data were perceived as generally relevant and not highly private. In addition to privacy and relevance, less common themes included **access to the data source** (“I don’t even know my family’s mental health history”, Georgie, 17 years old) and **curiosity about results** arising from analysis of particular data sources (“I search all weird stuff, so I thought it might be interesting to see if they have any links to anything”, Claire, 17).

### Data sharing between systems

The great majority of participants agreed with the health system accessing their school records (93.6%), and the school accessing health records (82.5%) for mental health predictive services. About a third of the participants indicated that sharing this information was **already common practice** (see **Fig3** for quotes). Over a quarter explicitly mentioned that they considered cross-sectoral sharing to be **beneficial** to them, and many referred to the information being **relevant to the (early) detection of mental health challenges**. The minority who refused to share data with school typically argued that mental health screening was **not the school’s responsibility** (“I feel like that’s more something that should happen in the medical, like the doctor world, that kind of thing. I don’t think it’s for school”, Pam, 17 years old).

**Fig3.**
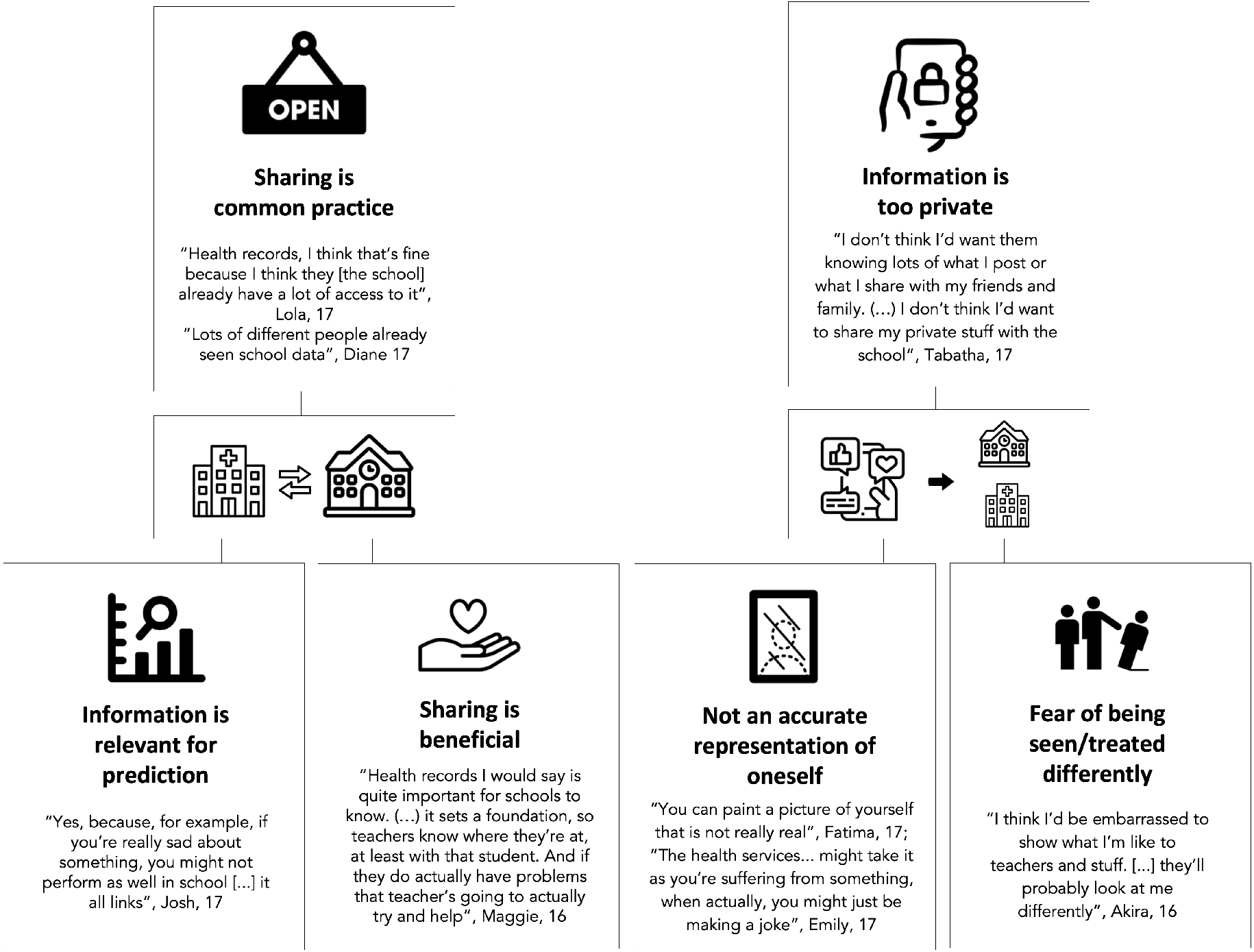
Main themes coded from participants’ arguments for sharing data between health and school systems (left) and for *not* sharing social media data with schools or health services to identify risk for mental health challenges

When it comes to sharing social media data, however, while most participants (67.1%) agreed to share that information with the health system, only 30% were willing to share it with schools. When discussing social media data sharing, most participants raised **privacy concerns** due to the personal nature of this information, and over a quarter indicated that **predictions based on social media data might not be accurate**, with some fearing being misinterpreted by an algorithm. A minority expressed concerns that they might be **seen or treated differently** by school staff specifically, should they know details about their personal life or mental health (see **Fig3** for quotes). Willingness to share was largely justified based on the participant **not posting information that is personal or secretive** through social media platforms (“Yes, I don’t mind. There’s nothing. Nothing there”, Miley, 17 years old), and a minority indicated the **information could be relevant** for the prediction (“Actually yes, because some people, when they’re depressed, they go on social media, and they do those posts like, I’m really depressed, or whatever”, Steve, 18 years old). Further benefits were rarely mentioned, and those willing to share often emphasised only feeling comfortable sharing data already set as public (e.g., open feed rather than direct messages).

## CONCLUSIONS

The aim of our study was to investigate young people’s interest in predictive testing services in psychiatry, and their preferences and attitudes regarding sharing biological, psychological, and behavioural data for a risk assessment. Using a combination of digital role-play and peer-led interviews, we found that most participants were interested in taking a predictive test, especially for anxiety, learning difficulties, and depression. Young people were motivated by the perceived relevance of mental health to themselves and their daily life; the opportunity to learn more about themselves; and the chance to engage in preventative measures. Participants were willing to share data sources they perceived as relevant for making an accurate prediction; however, they were reluctant to share certain types of digital data due to privacy concerns (e.g., text messages and phone conversations). With regards to cross-sectoral data sharing, participants were positive about sharing school records with the health system, and health records with schools; however, most were reluctant to share social media data with schools (but less reluctant to share these data with the health system).

Participants’ high interest in taking a predictive psychiatric test aligns with research showing that adolescents value learning about themselves and their health [23]. Participants’ differential interest in testing for specific conditions suggests a reasoned approach to healthcare decision-making. Notably, they were specifically motivated if they found the test personally relevant to them e.g., due to familial psychiatric history or emergent symptomatology. This is supported by prior research that documented high interest in learning about one’s risk for developing psychosis amongst clinical high-risk adolescents [20] as well as for major depressive disorder for those with familial experience of depression [27].

Whilst adolescents in our study valued the idea of predictive services in psychiatry, they were selective with regards to what data sources to share for the assessment. Specifically, participants demonstrated significant reluctance to share digital footprints, such as social media data (in comparison to psychosocial and biological data), because of *both* concerns around privacy and – for some data sources – a perceived lack of utility. This caution around data privacy in a digital context is mirrored by existing research demonstrating the high value attributed to digital privacy by adolescents [28]. Future research should investigate whether providing young people with reassurances that their data will be kept safe and private increases their willingness to share digital footprints.

Results also indicated that young people base their decisions to share certain types of data not only on its usefulness and privacy, but on the end recipient of the shared data (e.g., schools or health systems). These responses corroborate research highlighting the hesitancy of patients to share data when they have limited information or trust regarding its confidentiality [29]. Unless the reasons for young people’s hesitancy to share digital data with schools are addressed and carefully considered by educational settings, the promise of social media-based algorithms to act as “virtual gatekeepers” [30] will not be achieved.

### Methodological strengths and limitations

Our digital role-play engaged players in an immersive experience [25], thereby creating a model that gets closer to the actual use of services for mental health risk assessments. However, it is possible that different decisions would be made in a real-life setting or a different simulated scenario. The peer-to-peer interview format facilitated self-initiated discussion, akin to a real-life conversation between peers; by challenging each other, participants were able to engage in deeper ethical reflection. However, it is also possible that adolescents influenced each other’s responses and arrived at converging arguments. Finally, our sample was diverse but not nationally representative, and there might have been an overrepresentation of adolescents interested in mental health (who voluntarily signed up). Future research using different methods, and involving a larger, representative sample of UK adolescents will help ascertain the extent to which the current results are replicable and generalisable.

## CLINICAL IMPLICATIONS

Data from digital technologies such as wearables and smartphones hold potential for detecting mental health challenges at an early stage, and supporting early intervention and selective prevention in adolescent mental health. The demand for this technology has led to ‘function-creep’: the repurposing of software programmes to monitor young people’s mental health. Such re-purposing has been especially prevalent within the educational context, where, for example, software used primarily for homework submission integrated additional capabilities for mental health monitoring. Social media platforms have also integrated mental health monitoring features (e.g., Facebook monitoring suicide risk). In some cases, opting out of monitoring means opting out of the main service, leaving young people little choice on the use of their personal data.

As this study shows, young people see multiple benefits in predictive psychiatric services, but wish to have choice over which type of data is shared, with whom and for what purpose. Moreover, young people wish to know why different data sources are relevant for mental health prediction and how their privacy will be protected. Given young people’s reluctance to share digital footprints, providers of predictive psychiatric services must consider whether the relevance of different digital data sources for psychiatric assessments outweighs privacy concerns they might present to young people. Respecting adolescents’ need for privacy, transparency and choice in the age of digital phenotyping will be critical for responsible design and implementation of preventive psychiatric technologies and services in the future.

## Data Availability

Data are available upon request

https://osf.io/cwjx4/

## ACKNOWLEDGMENTS

We thank the NeurOX Young People’s Advisory Group and the BeGOOD Work Experience Team for their participation in the development of the role-play and interview guide. IS, GP and JL are supported by a Wellcome Trust Senior Investigator Award to IS (Grant 104825/Z/14/Z). IS is, in addition, supported by the NIHR Oxford Health Biomedical Research Centre under Grant IS-BRC-1215-20005 and the Wellcome Centre for Ethics and Humanities, which is supported by core funding from the Wellcome Trust under Grant 203132/Z/16/Z. The project was made possible by the Junior Researcher Programme (http://jrp.pscholars.org/).

## Contribution statement

GP and IS conceived the study, and all authors contributed to methodology design and e-tool development. GP, JL, KW, BW, LK, LG and SA carried out piloting and data collection and JL prepared the data for analysis. GP, AY, KW, BW, NT, LK, LG and SA analysed the data, interpreted the results, and contributed to the first draft of the manuscript. IS and JL critically revised the paper for important intellectual content. GP finalised the paper following co-authors’ review, and all authors approved the final version. IS is responsible for the overall content as guarantor.

## Footnotes

1 After completing the first part of the peer-led interview, participants completed a second role-play task and interview. Data from this additional section is beyond the scope of the present paper and therefore will not be reported.

